# Determinants of Patient Satisfaction in a Bangladeshi Public Hospital Outpatient Department: A Cross-Sectional Study

**DOI:** 10.1101/2025.08.01.25332704

**Authors:** Md. Rizwanul Karim, Shahnaz Akhter, Taslima Zannat, Tahmid Sajid

## Abstract

**Background:** Patient satisfaction is critical for healthcare utilization in resource-constrained settings like Bangladesh, yet evidence on outpatient satisfaction determinants remains limited. This study assessed perceived service quality and identified predictors of satisfaction in a public medical college hospital.

**Methods:** A cross-sectional study (March–May 2025) recruited 1,089 adult outpatients via systematic random sampling. Data were collected using validated Bengali instruments measuring perceived quality (24 items) and satisfaction (14 items). Binary logistic regression was used to identify significant predictors (p < 0.05). The model was validated using the area under the curve (AUC), pseudo-R², and classification accuracy. Results were reported as odds ratios with 95% confidence intervals. Data entry, editing, and analysis were performed using SPSS and Jamovi software.

**Results:** Participants (mean age 34±15.64 years; 51.5% male; 63.0% rural) reported positive physician interactions (e.g., 56.7% felt respected). Critical deficiencies included poor toilet cleanliness (75.5%), inadequate drinking water (67.6%), and medication shortages (30.6%). Female patients reported insufficient consultation time (17.5%). Regression analysis (AUC=0.92) revealed higher satisfaction among males (AOR=1.79, 95% CI: 1.02–3.16), suburban residents (vs. rural; AOR=2.01, 1.06–3.81), businesspersons (vs. students; AOR=2.89, 1.19–7.04), and those with prior positive experiences (AOR=1.50, 1.42–1.58). Upper-middle-class patients had lower satisfaction (AOR=0.40, 0.20–0.79). Positive perceptions of management and administration independently increased satisfaction (AOR=1.14 for both).

**Conclusion:** Patient satisfaction in a Bangladeshi public hospital outpatient department is influenced by multifaceted determinants: demographics, service experiences, and perceived quality. Key predictors of higher satisfaction included male gender, suburban residence, business occupation, positive prior experiences, and perceived administrative quality. Conversely, upper-middle-class status and female gender predicted lower satisfaction, indicating equity concerns. These findings necessitate urgent, targeted improvements in non-clinical areas (administration, resource logistics, facility environment) and gender-sensitive, equity-focused interventions to enhance satisfaction, trust, and service utilization.

**What is already known:** Patient satisfaction in LMIC public health settings is shaped by clinical interactions (e.g., provider competence) and non-clinical factors (e.g., waiting times, sanitation). Gender and socioeconomic disparities persist, but evidence rarely controls for prior experiences, risking confounding.

**What this study adds:** In 1,089 Bangladeshi OPD patients:

1. Quantified disparities: Males had 79% higher satisfaction than females (AOR=1.79); upper-middle-class patients had 60% lower satisfaction than lower-class peers (AOR=0.40).
2. Isolated current drivers: Controlling for prior satisfaction, management (ρ=0.28), and administration (ρ=0.22) independently predicted satisfaction.
3. Exposed systemic gaps: 75.5% reported poor sanitation; 17.5% of women received insufficient consultation time.

**Implications:**

- Research: Control for prior satisfaction in patient experience studies.
- Practice: Prioritize gender-transformative training and sanitation investments.
- Policy: Integrate patient feedback systems and equity standards for rural access

## Background

Patient satisfaction and perceived service quality are increasingly recognized as key indicators of health system performance, particularly in Low- and Middle-Income Countries (LMICs). These constructs play a crucial role not only in assessing care delivery but also in shaping patients’ health-seeking behavior, compliance with medical advice, and willingness to utilize public healthcare facilities again. In LMICs like Bangladesh, where government healthcare services are often provided free or at minimal cost, ensuring high patient satisfaction is essential to increase service utilization, build trust, and reduce dependence on out-of-pocket private services [1].

Numerous studies from South Asia and similar resource-constrained settings have emphasized that patient satisfaction is influenced by a complex interplay of factors, including the technical competence of healthcare providers, interpersonal communication, administrative efficiency, physical environment, and system-level organization. For instance, research from India and Nepal indicates that although public health facilities are often staffed by competent clinicians, deficiencies in hygiene, overcrowding, lack of information, and staff behavior contribute significantly to patient dissatisfaction, ultimately reducing the utilization of otherwise free services [2–4].

Several studies have also highlighted the importance of non-clinical dimensions of care, such as waiting time, staff courtesy, privacy, and sanitation, as essential determinants of patient satisfaction [1, 5]. While physicians’ technical capabilities were rated positively, dissatisfaction with hospital hygiene, waiting times, and staff responsiveness indicates that non-clinical services are key drivers of satisfaction. Moreover, a lack of information about diagnostic procedures, medication availability, and monitoring during treatment is a known deterrent to public healthcare engagement [6]. These gaps in perceived quality of care, regardless of cost, can lead patients to seek expensive private alternatives, resulting in unnecessary financial burdens and underutilization of otherwise accessible public healthcare services [7]. These dimensions are particularly relevant in outpatient departments (OPDs), where care is episodic and often fast-paced. A multi-country analysis by the WHO South-East Asia office emphasized the role of patient-centered communication and dignity in shaping overall satisfaction and the likelihood of returning to a facility [8]. Patients who view services as unresponsive or disrespectful may avoid future visits, even if the services are accessible and free. Patient satisfaction strongly predicts future service utilization. Governments should thus integrate satisfaction monitoring into routine quality assurance. Feedback-driven reforms can align services with patient expectations, improving trust and care continuity [9–10].

Furthermore, socioeconomic and geographic disparities influence how patients perceive and interact with the healthcare system. Patients from rural areas and lower-income groups were more likely to express dissatisfaction with government health services due to perceived neglect and discrimination [11]. In South Asia, similar disparities are observed, such that suburban patients often receive better interpersonal care compared to rural counterparts, despite standardized protocols within public hospitals [12]. Importantly, gender-sensitive service delivery remains a significant gap in Public-sector healthcare across many LMICs. Evidence from Pakistan and India demonstrates that female patients are more likely to face inadequate consultation time, less privacy, and poorer communication compared to male patients [13–15]. These deficiencies contribute to unmet health needs and underutilization of maternal and reproductive health services, even when such services are provided for free. Addressing these gender-based disparities is crucial for advancing equity and improving health outcomes among underserved populations.

### Research Gap

While Bangladesh’s public hospitals face known quality challenges, evidence gaps persist in outpatient satisfaction determinants, particularly regarding:

(1) Validated quantification of non-clinical service dimensions (administrative efficiency, gender sensitivity),
(2) Intersectional disparities (rural-urban/socioeconomic/gender gradients), and
(3) Systemic predictors of satisfaction in resource-constrained tertiary settings. Existing studies rely on non-representative samples or inpatient focus, limiting actionable insights for equity-oriented reforms in high-volume OPDs.

### Objective

The primary objective of the study is to assess perceived service quality and patient satisfaction in the outpatient department of a public medical college hospital in Bangladesh. The study also aims to examine how demographic, institutional, and service-related factors influence satisfaction outcomes, using standardized and validated assessment tools.

## Methodology

### Study Design and Setting

This cross-sectional study was conducted from March to May 2025 in the outpatient department of Shaheed M. Monsur Ali Medical College (SMMAMC), Bangladesh. The tertiary facility serves approximately 50,000 annual patients from Sirajganj district, where residents predominantly belong to lower and lower-middle socioeconomic groups. This setting exemplifies Bangladesh’s public medical colleges, characterized by high patient volumes operating within constrained resource environments. The study focused on patients who received outpatient medical services but were not admitted to the inpatient department.

### Study Population and Sampling Technique

The study population consisted of patients attending the OPDs during the study period. Participants were selected using a systematic random sampling technique. Every 6th patient attending the outpatient department was approached for participation after applying the inclusion and exclusion criteria.

### Inclusion and Exclusion Criteria

Patients were included in the study if they had received outpatient services at the hospital and provided written informed consent. Individuals were excluded if they were seriously ill, had a major psychiatric illness, were under the age of 18, or declined to participate.

### Sample Size Determination & sampling

The minimum sample size was calculated using an assumed patient satisfaction prevalence of 51% in Bangladeshi public medical college hospitals [16]. Parameters included a 95% confidence level (Z_α/2_= 1.96), 90% statistical power (Z_β_= 1.28), a 3% margin of error, and a design effect of 1. Using *Ausvet Epitools software*, the initial calculation yielded 1,067 participants. To compensate for potential non-response, the estimated sample was inflated by 5%, resulting in a final requirement of 1,120 participants. This ensured adequate precision to detect significant associations while maintaining robustness against attrition.

Over eight weeks (32 working days), systematically selected participants underwent interviews in the outpatient department (OPD), which averaged 200 ± 50 daily patient visits. The sampling interval was based on an estimated 6,000–8,000 patient visits over 8 weeks, with a target of 1,120 participants. Accordingly, every 6th eligible patient was systematically chosen and invited to join. Using systematic random sampling, every 6th eligible patient was invited to participate. A new random starting point (range: 1–6) was designated daily to mitigate selection bias. If a patient declined or was ineligible, the next suitable candidate was enrolled, maintaining a daily recruitment target of ∼35 participants. Administrative approvals facilitated seamless implementation. From an initial target of 1,120 respondents, 1,089 complete responses were retained after data cleaning for final analysis.

### Study Instrument and Variables

Data were collected using a semi-structured, interviewer-administered questionnaire. The questionnaire captured a wide range of information, including:

* Sociodemographic characteristics
* Disease and service-seeking characteristics
* Perceived quality of services
* Perceived present and past satisfaction with outpatient services

Perceived quality and satisfaction were assessed using validated Bengali versions of two measurement tools. While previously used in community clinic settings, these tools were believed to be contextually, structurally, and culturally appropriate for use in the outpatient department (OPD) settings of public hospitals. Each tool was made up of 24 items, rated on a Likert scale, and demonstrated strong psychometric properties established in previous validation studies [1]. The perceived quality scale’s four-component structure explained 60.7% of the variance with high internal consistency (α = 0.89; subscale α = 0.74–0.90). The satisfaction scale explained 71.16% of the variance, showing excellent internal consistency (α = 0.97; subscale α = 0.86–0.94) [1].

### Ethical Considerations

This study was conducted in full compliance with the Declaration of Helsinki and received ethical approval from the Shaheed M. Monsur Ali Medical College IRB (Memo No. IEC/2025/087, Clearance Certificate: IEC-CERT-2025-087). Before participation, all individuals were thoroughly informed about the study’s objectives, procedures, and their rights, including the freedom to participate voluntarily and withdraw at any time without consequence. Written informed consent was obtained from each participant. Strict measures were taken to ensure confidentiality and anonymity—no personally identifiable information was collected, all data were securely managed, and results are reported in aggregate form exclusively for academic and public health purposes.

### Statistical Analysis

Data analysis was conducted using SPSS version 23 and Jamovi 2.6.44.0. Satisfaction scores were dichotomized at the median value of 42 to mitigate arbitrary thresholds and balance group sizes for robust comparisons. This approach addressed non-normal data distributions with clustered extreme responses while maximizing statistical power for logistic regression. The median cutoff also aligned with the scale’s conceptual midpoint: as each of the 14 items used a 5-point Likert scale (where 3 = ‘Satisfied’), a score of 42 represented the theoretical neutral point between satisfaction and dissatisfaction. Descriptive statistics (frequencies, percentages) were used to summarize the responses related to perceived service quality and satisfaction. Partial correlation was performed to control for potential confounding from past-year satisfaction scores, allowing for clearer interpretation of associations among service domains (e.g., skill, management, administration, environment) and satisfaction outcomes. Spearman’s rank-order correlation was applied due to the non-parametric nature of the data. For inferential analysis, binary logistic regression was used to identify significant predictors of patient satisfaction. All variables with theoretical or empirical relevance were included in the multivariable model. Model adequacy, multicollinearity, classification accuracy, sensitivity, specificity, receiver operating characteristic curve, and area under the curve (AUC) were computed to evaluate model performance. Statistical significance was set at p < 0.05 for all tests.

## Results

A random sample of 1,089 patients visiting the Outpatient Department (OPD) was interviewed to assess their perceptions of service quality and overall satisfaction. Among these, 33% (n=361) reported having been ill in the previous six months. The respondents had an average age of 34 years (SD = 15.64), with 51.5% (n = 561) being male and 48.5% (n = 528) female. The majority identified as Muslim (95%, n =1036) and were married (73.6%, n = 802). Most lived in villages (63%, n = 681), while 11% (n = 120) were from suburban areas. Housewives were the largest occupational group (33%, n = 359), followed by students (24.8%, n = 270), day laborers (11.8%, n = 128), and business owners (10%, n = 105). Socioeconomic status included 41.8% (n=455) upper-middle income, 34.4% (n=375) lower-middle income, and 17.9% (n=195) classified as poor. Health-risk behaviors involved tobacco smoking (13%, n =142) and smokeless tobacco use (12%, n=131). Before visiting the public hospital, 34.7% (n = 378) sought care from pharmacies and 15.2% (n=165) from village doctors. The average health expenditure was 2,612 Bangladeshi Taka (BDT) (SD = 1,900), and the typical travel time to the hospital OPD was 54 minutes (SD = 40).

Patient perceptions of OPD service quality showed notable differences across different areas. Interpersonal aspects of care (Skill, competence & communication) received the highest ratings: most patients reported good levels of respect (56.7%), sympathy (58.1%), kindness (49.4%), and honesty (54.1%) from physicians, while many felt comfortable sharing information (50.0% rated as good). However, views on managerial and administrative factors were less positive. Only 43.9% rated physicians’ ability to give disease advice as good, and 42.3% believed examinations were thorough. Time constraints emerged as a major issue, with 22.4% perceiving physicians as rushed and 17.5% reporting they spent insufficient time with female patients. Resource shortages and follow-up gaps were also evident: 30.6% experienced medication shortages or interruptions, 36.1% felt follow-up monitoring was inadequate, and 31.8% noted a shortage of ancillary staff. Sanitation (Physical facility amenities) issues were especially severe, with hygiene being a significant concern—75.5% rated toilet cleanliness as not good, and 67.6% reported poor access to drinking water. In contrast, hospital organization (58.9% overall) and general environmental cleanliness (65.2% overall) received more moderate ratings. These findings highlight a divide between strong patient-physician relationships and systemic challenges related to time management, resource availability, and environmental conditions. hygiene. [Table 1].

**Table 1:** Patient Perceptions of Hospital Outpatient Service Quality Across Skill, Management, Administration, and Environment Domains (N = 1,089)

| Questionnaire about Perceived Quality | Not good<br>N (%) | Average<br>N (%) | Good<br>N (%) |
| --- | --- | --- | --- |
| 1. In your opinion, how capable is the doctor to advise you about the diseases? | 107 (9.8) | 504 (46.3) | 478 (43.9) |
| 2. How much respect does the doctor have for you? | 41(3.8) | 430 (39.5) | 618 (56.7) |
| 3. What do you think about the capability of the doctor in determining the disease? | 99 (9.1) | 432 (39.7) | 558 (51.2) |
| 4. Do you feel comfortable disclosing your information to the doctor? | 76 (6.9) | 469 (43.1) | 544 (50.0) |
| 5. How thoroughly did the doctor examine you? | 106 (9.7) | 522 (47.9) | 461 (42.3) |
| 6. How sympathetic is the doctor to you? | 50 (4.6) | 406 (37.3) | 633 (58.1) |
| 7. Did you find the doctor honest? | 80 (7.3) | 420 (38.6) | 589 (54.1) |
| 8. How much time does the doctor spend during the treatment of female patients? | 191(17.5) | 532 (48.9) | 366 (33.6) |
| 9. Is the doctor in a hurry while consulting with the patient? | 244 (22.4) | 528 (48.5) | 317 (29.1) |
| 10. Did the doctor seem kind? | 92 (8.4) | 459 (42.1) | 538 (49.4) |
| 11. Have you been properly informed about the diagnostic tests & procedures? | 187 (17.2) | 523 (48.0) | 379 (34.8) |
| 12. How effective is their prescription? | 277 (25.4) | 396 (36.4) | 416 (38.2) |
| 13. Did the hospital provide you with enough medication? | 333 (30.6) | 553 (50.8) | 203 (18.6) |
| 14. Were you properly observed and monitored (follow-up)? | 393 (36.1) | 526 (48.3) | 170 (15.6) |
| 15. How organized were the overall hospital healthcare facilities? | 96 (8.8) | 641 (58.9) | 352 (32.3) |
| 16. Did the doctor spend enough time explaining the disease treatment? | 192 (17.6) | 551 (50.6) | 346 (31.8) |
| 17. Has your health improved after treatment? | 194 (17.8) | 460 (42.2) | 435 (39.9) |
| 18. Were adequate support (ancillary) staff present in the hospital? | 347 (31.8) | 554 (50.9) | 188 (17.3) |
| 19. How was the Hospital Environment (supportive, ensure privacy)? | 251 (23.0) | 567 (52.1) | 271 (24.9) |
| 20. Whether adequate Healthcare provider (Doctor, Nurse, Medical assistant, etc.).? | 265 (24.3) | 566 (52.0) | 258 (23.7) |
| 21. How is the time management of the hospital staff? . | 305 (28.0) | 476 (43.7) | 308 (28.3) |
| 22. What do you think about the hygiene of the hospital toilets? | 822 (75.5) | 232 (21.3) | 35 (3.2) |
| 23. How is the availability of a clean water supply in the hospital? | 736 (67.6) | 301 (27.6) | 52 (4.8) |
| 24. How clean is the hospital environment in your view? | 185 (17.0) | 710 (65.2) | 194 (17.8) |

*The categories “Do not know” and “Not good” were combined into a single category, “Not good,” to facilitate* *clearer reporting and enhance interpretability.*

In our study, patient satisfaction data, particularly on women’s experiences with OPD services, showed mostly positive feedback about communication, provider interactions, and care delivery.

A significant majority of respondents reported satisfaction with staff courtesy (81.7%), the ability to maintain privacy during consultations (83.7%), and clarity in communication (77.4%). Notably, 86.3% of women expressed satisfaction with how seriously their health concerns were addressed. However, there were lower satisfaction rates concerning system organization. Only 57.9% of respondents were satisfied with updates on waiting times, and 61.7% felt the same about the flexibility of appointment scheduling. These insights suggest that while interpersonal care was valued, there is a need for improvements in service management and administration.

Last year’s experiences from the OPD department showed high satisfaction levels in trust towards health professionals (78.3%) and knowledge of women’s health issues (77.4%). Lower satisfaction levels were found in the prevention focus (57.8%) and emotional health interest (62.8%). This indicates a need for improved preventive care and emotional support. [Table 2].

**Table 2:**
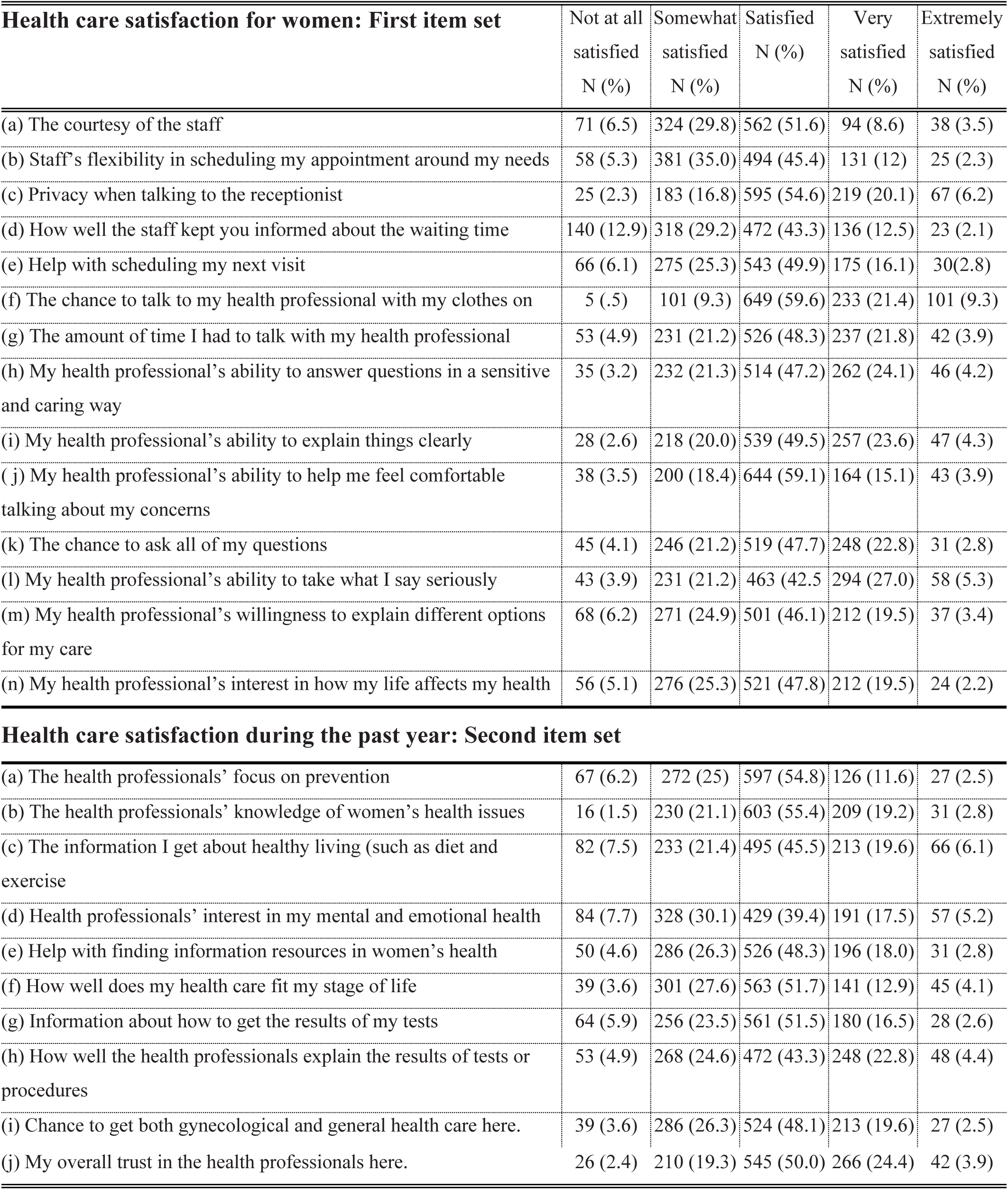
Patient satisfaction with OPD services [N = 1089].

A partial correlation analysis was conducted to examine how perceived quality dimensions relate to overall OPD satisfaction, while controlling for past satisfaction to account for previous experiences. The satisfaction score showed a weak yet significant positive correlation with the Skill score (Spearman’s rho = 0.26, p = 2.17e-18) and the Management score (rho = 0.28, p = 1.44e-20). Stronger correlations were observed between the Skill score and the Management score (rho = 0.52, p = 3.09e-76), as well as between the Management score and the Administration score (rho = 0.53, p = 8.01e-81). The Administration score also correlated moderately with the Skill score (rho = 0.41, p = 2.84e-45) and weakly with the Environment score (rho = 0.27, p = 2.53e-19). The Environment score exhibited the weakest but still significant correlations with the Satisfaction score (rho = 0.13, p = 3.05e-5) and the Skill score (rho = 0.16, p = 2.56e-7). All reported correlations were statistically significant (p < .001), indicating robust relationships among the variables after accounting for past satisfaction. [Table 3].

**Table 3:**
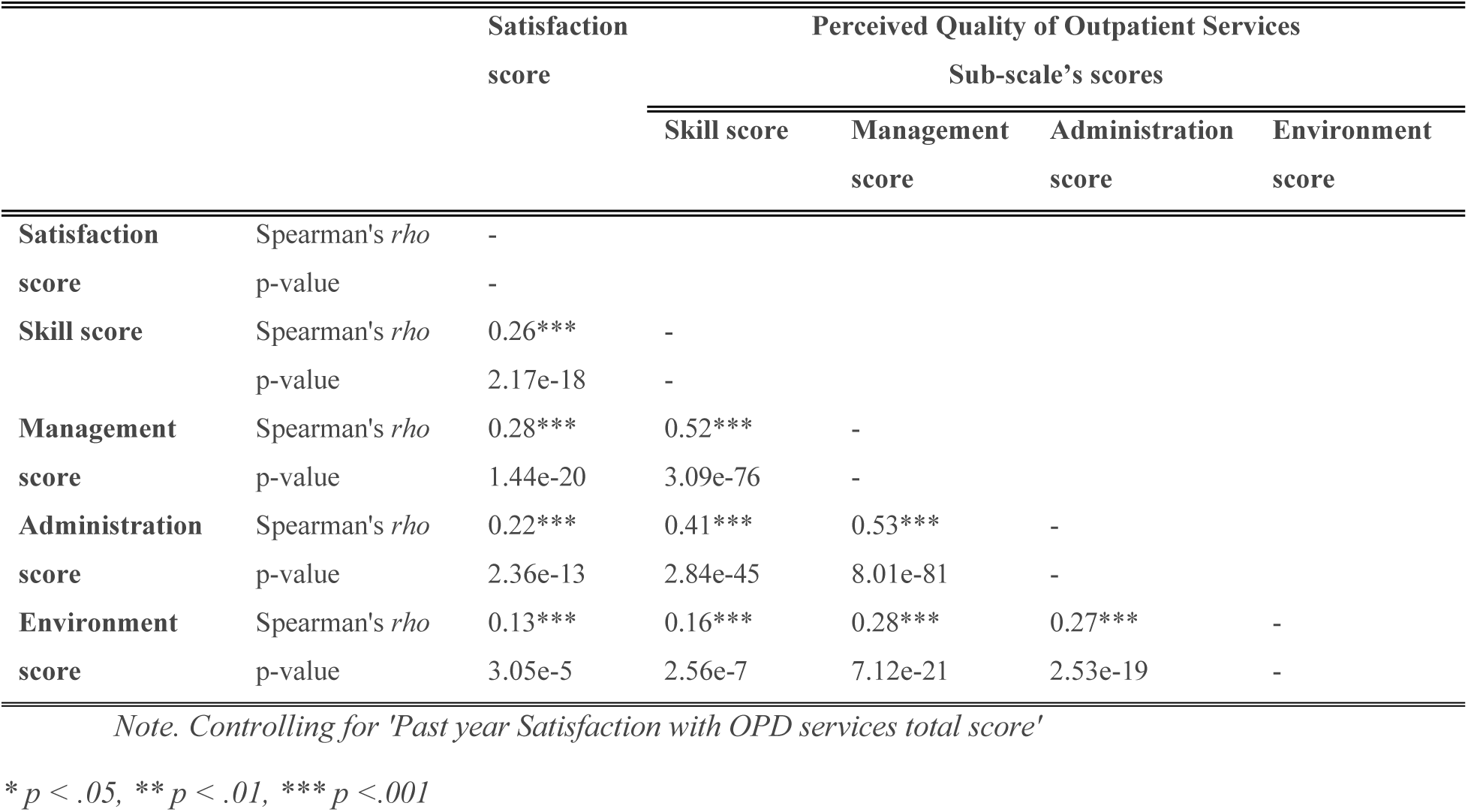
Partial correlation was used to assess the relationship between perceived quality dimensions and OPD satisfaction, controlling for prior satisfaction.

|  |  | Satisfaction | Perceived Quality of Outpatient Services |  |  |  |
| --- | --- | --- | --- | --- | --- | --- |
|  |  | score | Sub-scale's scores |  |  |  |
|  |  |  | Skill score | Management | Administration | Environment |
|  |  |  |  | score | score | score |
| Satisfaction score | Spearman's *rho* | - |  |  |  |  |
|  | p-value | - |  |  |  |  |
| Skill score | Spearman's *rho* | 0.26*** | - |  |  |  |
|  | p-value | 2.17e-18 | - |  |  |  |
| Management score | Spearman's *rho* | 0.28*** | 0.52*** | - |  |  |
|  | p-value | 1.44e-20 | 3.09e-76 | - |  |  |
| Administration score | Spearman's *rho* | 0.22*** | 0.41*** | 0.53*** | - |  |
|  | p-value | 2.36e-13 | 2.84e-45 | 8.01e-81 | - |  |
| Environment score | Spearman's *rho* | 0.13*** | 0.16*** | 0.28*** | 0.27*** | - |
|  | p-value | 3.05e-5 | 2.56e-7 | 7.12e-21 | 2.53e-19 | - |

*Note. Controlling for 'Past year Satisfaction with OPD services total score'*
\* $p < .05$ , \*\* $p < .01$ , \*\*\* $p < .001$

### Predictors of Patient Satisfaction

A binary logistic regression analysis was employed to identify predictors of patient satisfaction, which was dichotomized based on a median cut-off score of 42 from the 14-item Patients’ Satisfaction Scale (First items). Patients scoring below 42 were classified as “not satisfied,” while those scoring 42 or above were categorized as “satisfied.“

The regression model demonstrated excellent predictive performance (AUC = 0.92) and goodness-of-fit (χ² = 717.77, *p* < 0.001). Key diagnostics, including strong pseudo-R² values (McFadden’s = 0.47; Nagelkerke’s = 0.64), favorable information criteria (AIC/BIC = 882.71/1117.38), and absence of multicollinearity (max VIF = 1.62)collectively support model robustness. Classification accuracy was high (85%), with balanced sensitivity (84%) and specificity (85%). Full diagnostics are reported in Table 4.

**Table 4:**
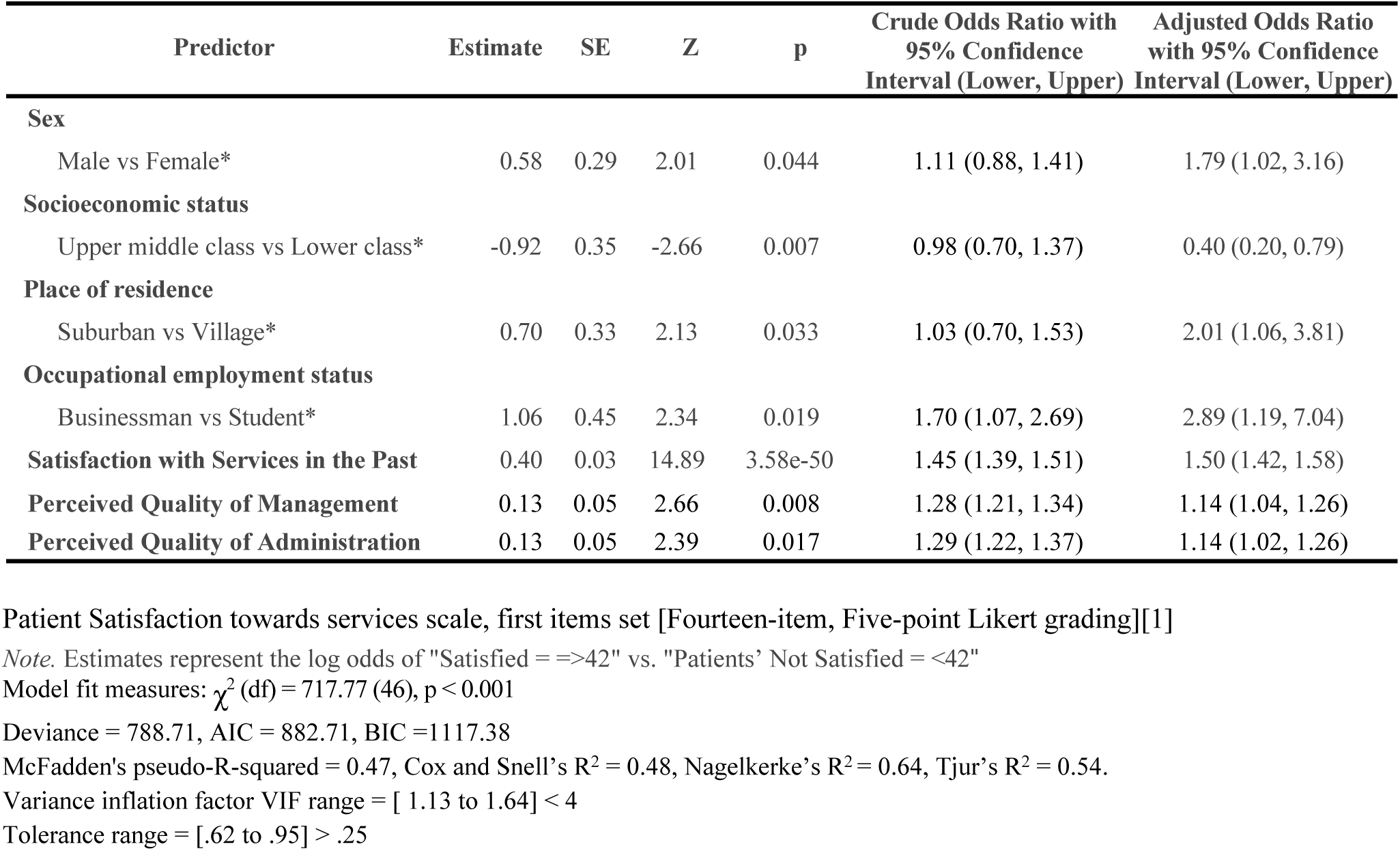
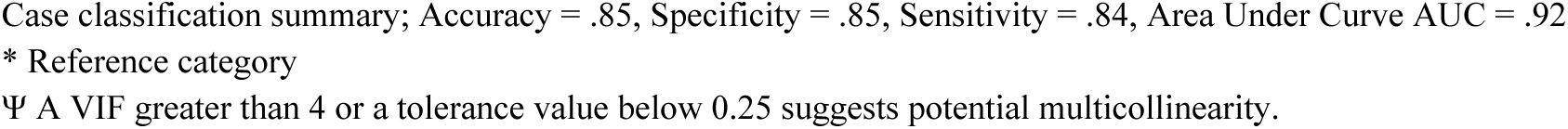
Predictors of Patients’ Satisfaction in the Sample Population [N = 1089].

| Predictor | Estimate | SE | Z | p | Crude Odds Ratio with 95% Confidence Interval (Lower, Upper) | Adjusted Odds Ratio with 95% Confidence Interval (Lower, Upper) |
| --- | --- | --- | --- | --- | --- | --- |
| **Sex** |  |  |  |  |  |  |
| Male vs Female* | 0.58 | 0.29 | 2.01 | 0.044 | 1.11 (0.88, 1.41) | 1.79 (1.02, 3.16) |
| **Socioeconomic status** |  |  |  |  |  |  |
| Upper middle class vs Lower class* | -0.92 | 0.35 | -2.66 | 0.007 | 0.98 (0.70, 1.37) | 0.40 (0.20, 0.79) |
| **Place of residence** |  |  |  |  |  |  |
| Suburban vs Village* | 0.70 | 0.33 | 2.13 | 0.033 | 1.03 (0.70, 1.53) | 2.01 (1.06, 3.81) |
| **Occupational employment status** |  |  |  |  |  |  |
| Businessman vs Student* | 1.06 | 0.45 | 2.34 | 0.019 | 1.70 (1.07, 2.69) | 2.89 (1.19, 7.04) |
| **Satisfaction with Services in the Past** | 0.40 | 0.03 | 14.89 | 3.58e-50 | 1.45 (1.39, 1.51) | 1.50 (1.42, 1.58) |
| **Perceived Quality of Management** | 0.13 | 0.05 | 2.66 | 0.008 | 1.28 (1.21, 1.34) | 1.14 (1.04, 1.26) |
| **Perceived Quality of Administration** | 0.13 | 0.05 | 2.39 | 0.017 | 1.29 (1.22, 1.37) | 1.14 (1.02, 1.26) |

Patient Satisfaction towards services scale, first items set [Fourteen-item, Five-point Likert grading][1]
Note. Estimates represent the log odds of "Satisfied = $\geq 4$ " vs. "Patients' Not Satisfied = $< 4$ "
Model fit measures: $\chi^2$ (df) = 717.77 (46), $p < 0.001$
Deviance = 788.71, AIC = 882.71, BIC = 1117.38
McFadden's pseudo-R-squared = 0.47, Cox and Snell's $R^2 = 0.48$ , Nagelkerke's $R^2 = 0.64$ , Tjur's $R^2 = 0.54$ .
Variance inflation factor VIF range = [ 1.13 to 1.64] $< 4$
Tolerance range = [.62 to .95] $> .25$
Case classification summary; Accuracy = .85, Specificity = .85, Sensitivity = .84, Area Under Curve AUC = .92
\* Reference category

The logistic regression analysis identified multiple significant predictors of patient satisfaction (defined as scores ≥42). Male patients demonstrated significantly higher satisfaction levels than female patients (AOR = 1.79, 95% CI [1.02, 3.16], p = 0.044). Conversely, patients from upper-middle-class backgrounds reported substantially lower satisfaction compared to their lower-class counterparts (AOR = 0.40, 95% CI [0.20, 0.79], p = 0.008). Regarding place of residence and occupational status, suburban residents exhibited significantly higher satisfaction levels than their rural counterparts (AOR = 2.01, 95% CI [1.06, 3.81], p = 0.033). Similarly, patients engaged in business reported nearly threefold greater satisfaction compared to students (AOR = 2.89, 95% CI [1.19, 7.04], p = 0.019). Furthermore, patients who reported higher satisfaction with prior healthcare services demonstrated significantly greater current satisfaction (AOR = 1.50, 95% CI [1.42, 1.58], p < 0.001). Positive perceptions of both management quality (AOR = 1.14, 95% CI [1.04, 1.26], p = 0.008) and administrative efficiency (AOR = 1.14, 95% CI [1.02, 1.26], p = 0.017) were also independently associated with elevated satisfaction levels. These results are visually summarized in the accompanying forest plot, highlighting the magnitude and precision of each predictor’s effect. These findings underscore the multifaceted nature of patient satisfaction, which encompasses demographic characteristics, prior experiences, and perceptions of healthcare services quality. [Table 4], [Figure 1].

**Figure 1.**
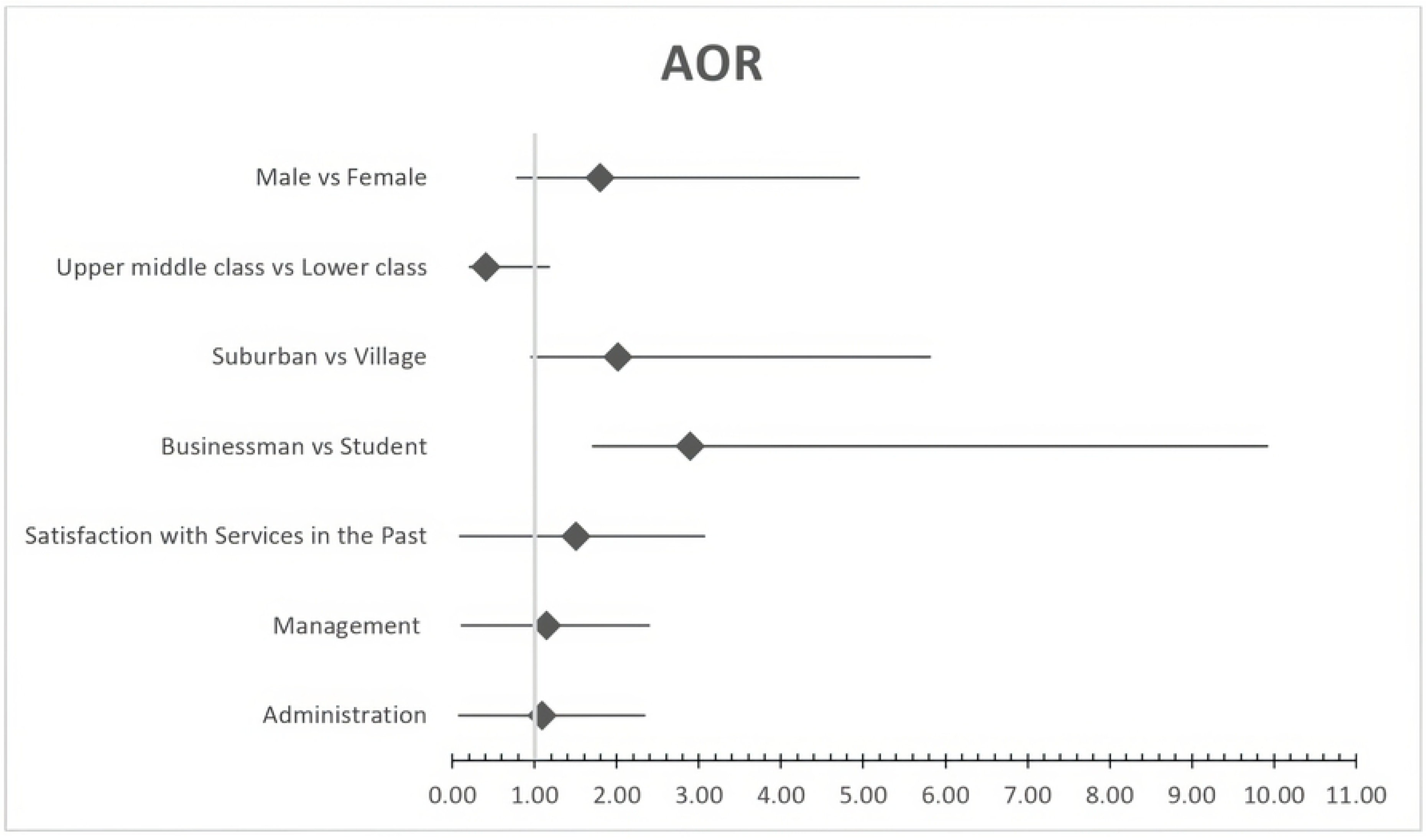
Predictors of Outpatient Satisfaction: Forest Plot of Adjusted Odds Ratios (AOR) from Binary Logistic Regression

## Discussion

This cross-sectional study offers valuable insights into patient satisfaction with outpatient services at a peripheral public medical college hospital in Bangladesh. Male gender, suburban residence, business occupation, positive prior healthcare experiences, and favorable perceptions of management quality and administrative efficiency independently increase outpatient care utilization satisfaction. Conversely, upper-middle-class socioeconomic status significantly decreases satisfaction. Therefore, patient satisfaction in Bangladesh’s resource-limited public hospital settings is affected by various factors, including demographic, experiential, and institutional aspects.

Our finding that male patients reported 79% higher satisfaction than females (AOR = 1.79) resonates with gendered care gaps across South Asia, where systemic biases limit consultation time and privacy for women [14,15]. This underscores the imperative for gender-transformative training—not merely ‘sensitive’ approaches—to dismantle entrenched inequities. Notably, upper-middle-class patients expressed significantly lower satisfaction than lower-income counterparts, aligning with recent regional analyses [17]. This may reflect heightened expectations arising from greater health system familiarity and prior exposure to private healthcare services, despite socioeconomic advantages. Similarly, suburban residents reported higher satisfaction than rural patients, corroborating studies from Nepal and Sri Lanka [12,14,18] that identify perceived inefficiency and disrespect in rural public healthcare delivery.

The study also found that business professionals and employed individuals had higher odds of satisfaction than students. This may reflect different expectations and thresholds of acceptability among various employment groups, as students may be more sensitive to delays or less empowered to demand respectful treatment. Comparable results from a Sri Lankan cross-sectional study suggested that occupational status influences health literacy and perceived entitlement, thereby shaping satisfaction [19].

Our findings indicated that although clinical care, particularly physician competence and respectful communication, was positively rated, notable dissatisfaction stemmed from managerial shortcomings such as poor information flow, inadequate medicine supply, staff shortages, and inefficient time management. This paradox is further exacerbated by the fact that, while physician competence and empathy were praised, systemic failures in sanitation, resource allocation, and gender-sensitive care undermined patient satisfaction. This misalignment mirrors trends across South Asia, where underfunded public systems struggle to meet non-clinical expectations [2–4,20].

Key administrative factors, including adequate staffing, staff behavior, protection of patient privacy, timely service delivery, and effective communication, were identified as strong predictors of patient satisfaction [1, 5, 21]. This aligns with WHO recommendations on people-centered care, which emphasize the importance of dignity and responsiveness in enhancing patient experiences and promoting greater utilization of health services [22]. There is a positive correlation between the quality of management and administration and overall patient satisfaction. Previous studies conducted in Bangladesh and India have shown that factors such as cleanliness of facilities, effective queue management, and courteous staff behavior play a crucial role in establishing patient trust. This is particularly important in outpatient departments, where patient interactions are often brief and infrequent [3–5].

Interestingly, patients who previously had positive experiences with the hospital’s OPD services were significantly more likely to report satisfaction again, illustrating the importance of perceived consistency in care. These prior positive experiences have been previously observed in LMICs, where even minor negative experiences can discourage future use of services and lead patients to opt for more expensive private alternatives [5, 23]. Given these results and patient involvement in service improvement, healthcare institutions should incorporate satisfaction monitoring as a core component of their quality assurance frameworks [24].

These findings indicate important implications for policy and practice. First, interventions should focus on reforming managerial and administrative processes. Key areas to address include the availability and timeliness of services, the supply of medications, monitoring procedures, patient privacy, waiting times, the allocation of time per patient, and staff responsiveness. Secondly, gender-sensitive training for healthcare workers should be arranged to minimize the disparity in satisfaction experienced by female patients. Thirdly, rural outreach and equity-focused quality improvements are necessary to bridge the suburban-rural-perception gap. Finally, feedback from patient satisfaction tools should be institutionalized to guide adaptive service delivery and resource allocation.

Despite methodological strengths, including a large sample, validated instruments, and advanced statistical analyses, this study has three principal limitations. The cross-sectional design precludes causal inference, while single-center sampling restricts generalizability to similar resource-constrained tertiary settings. Interviewer-administered Likert scales can introduce response biases, particularly central tendency and social desirability effects, where participants may adjust their responses to align with perceived norms. Although anonymity mitigated social desirability bias, residual inflation of satisfaction metrics remains possible.

### Policy & Practice Implications

→ Mandate gender-equitable consultation times and invest urgently in sanitation/medication supply chains.
→ Implement real-time patient feedback systems to drive resource allocation and equity-focused reforms.

### Conclusion& recommendations

This study concludes that patient satisfaction in the outpatient department of a public medical college hospital in Bangladesh is shaped by a complex interplay of demographic characteristics, patient experiences, and perceived service quality. While interpersonal aspects of physician care (skill, communication) were generally rated positively, significant issues in administrative efficiency, resource availability (especially medicines), time management, and sanitation (notably hygiene and water access) were major sources of dissatisfaction. Key predictors of higher satisfaction included male gender, suburban residence, business occupation, positive prior experiences, and favorable perceptions of management and administrative quality. Conversely, upper-middle-class status and female gender predicted lower satisfaction, highlighting concerns about equity. These findings emphasize the urgent need for targeted improvements in non-clinical service areas (administration, resource logistics, environment) and gender-sensitive, equity-focused interventions to improve patient satisfaction, trust, and utilization of public outpatient services in Bangladesh. Establishing patient feedback systems is essential for driving evidence-based, patient-centered quality improvements.

## Acknowledgments

Shaheed M. Monsur Ali Medical College Hospital, Directorate General of Medical Education.

## Availability of data and materials

The data that support the findings of this study are openly available in figshare at Karim, Md Rizwanul (2025). Determinants of Patient Satisfaction in a Bangladeshi Public Hospital Outpatient Department: A Cross-Sectional Study. figshare. Dataset. https://doi.org/10.6084/m9.figshare.29474936.v1

